# The Promise of mRNA Cancer Vaccines: Potential Lives Saved and Economic Value in the U.S.

**DOI:** 10.1101/2025.09.27.25336817

**Authors:** Chad R. Wells, Abhishek Pandey, Carolyn Bawden, Bilori Bilori, Ye Yang, Lilia Potter-Schwartz, Ayaz Lamia, Meagan C. Fitzpatrick, Alison P. Galvani

**Author notes:** **Corresponding authors:** Alison P. Galvani.

## Abstract

**Background:** The U.S. Department of Health and Human Services recently announced plans to curtail investment in messenger RNA (mRNA) vaccine development, despite the central role played by the platform in preventing millions of deaths during the COVID-19 pandemic. Beyond infectious diseases, mRNA vaccines are showing promise in oncology, where early-phase clinical trials report meaningful improvements in overall and recurrence-free survival. Evaluating the potential public health and economic value of these therapies is critical for informing funding decisions.

**Methods:** We reviewed ongoing mRNA cancer vaccine clinical trials and extracted available survival outcomes. To project potential impact of mRNA vaccination on overall survival, we combined trial-based improvements in survival with incidence and demographic-adjusted survival rates from the Surveillance, Epidemiology, and End Results (SEER) program of National Cancer Institute. A logistic regression framework estimated one- and three-year survival gains. We then applied the Value of a Statistical Life Year (VSLY, $604,000; 3% discount rate) provided by the U.S. Department of Health and Human Services to quantify the economic implications of forgoing mRNA investment.

**Results:** In a single annual U.S. cohort of patients newly diagnosed with non-small cell lung cancer, pancreatic cancer, renal cell carcinoma, or melanoma, mRNA vaccination could potentially avert approximately 49,000 deaths within three years of diagnosis. These projected survival gains translate to an estimated economic value of $75 billion.

**Conclusions:** Our findings underscore the substantial public health opportunity provided by mRNA cancer vaccines. Curtailing federal investment risks forfeiting these benefits, while sustained support could accelerate clinical translation and preserve infrastructure essential for future pandemic preparedness.

## Introduction

In August 2025, the U.S. Department of Health and Human Services announced it would cease federal investment in messenger RNA (mRNA) vaccine development, with the Biomedical Advanced Research and Development Authority (BARDA) cancelling more than $500 million in ongoing contracts.^1^ This decision comes at a pivotal moment for the field. While public funding was instrumental in accelerating mRNA vaccine development for COVID-19 that led to prevention of millions of deaths and saving hundreds of billions in healthcare costs,^1,2^ the withdrawal of support risks undermining progress in transformative oncology treatments.

Messenger RNA vaccines have emerged as one of the most powerful biomedical innovations of the past decade.^2,3^ Their unique advantages over traditional vaccine technologies include rapid design, scalability, and adaptability to both infectious and non-infectious diseases.^3,4^ Unlike conventional vaccines that rely on attenuated or inactivated pathogens, mRNA vaccines deliver genetic instructions that enable the body to produce antigens internally, eliciting strong immune responses with remarkable speed and flexibility.^2^ The success of COVID-19 vaccines highlighted not only their capacity to avert large-scale mortality and morbidity but also their unparalleled economic return on investment.

Building on this foundation, mRNA vaccines are increasingly being investigated for cancer treatment.^5,6^ Early results indicate treatment with mRNA vaccines can provide substantial improvements in overall survival, recurrence-free survival, and progression-free survival when compared to standard therapies.^7,8^ However, recent curtailing of federal investment at this juncture jeopardizes not only the translation of ongoing trials into approved therapies but also the infrastructure required to sustain rapid vaccine design and manufacturing capacity.^9,10^

In the present study, we evaluate the potential public health and economic benefits of mRNA cancer vaccines using preliminary trial results and national cancer data.^6,11^ Specifically, we reviewed active U.S.-based trials, modeled potential survival gains for selected cancers, and quantified the associated economic value of continued investment in this platform.

## Methods

We estimated the potential public-health and economic benefits of mRNA cancer vaccines by first identifying mRNA vaccine clinical trials and then extracting and harmonizing survival outcomes from these studies. We then used a logistic regression approach to estimate the survival benefit of potential mRNA vaccination compared to established treatments for four cancers with promising results. Next, we projected deaths averted in national incidence cohorts and computed economic cost averted by applying federal valuation benchmarks.

### Clinical trial identification

We systematically reviewed recent and historical literature reviews which mapped the current state of mRNA cancer vaccine research both in the United States and globally, and performed further research to find any clinical trials not included in these reviews. Trials were classified as “classic” mRNA or dendritic cell (DC) mRNA. Classic mRNA vaccines encode tumor-specific antigens into mRNA, which are then translated into proteins by the patients immune cells.^12^ In contrast, mRNA-DC vaccines involve the removal of DCs from the individual receiving treatment, which are then electroporated with tumor-attacking mRNA, and injected back into the body ^13^. These trials were screened to remove all trials not conducted and funded within the United States, as well as any trials that have been completed and thus not impacted by funding cuts. In all, 32 clinical trials were identified as mRNA cancer-vaccine trials currently being conducted within the United States, with 22 being classic mRNA vaccines and 10 being mRNA-DC vaccines. From these, we designated 11 trials as “promising” based on their trial phase (greater weight for Phase II/III), preliminary efficacy (characterized by improvements in overall survival rates, lowering of recurrence rates and delaying disease progression, and sponsor capacity to execute late-phase development and manufacturing.

We identified studies that had extractable overall survival (OS) data suitable for quantitative projection. These studies encompassed four cancers: non-small cell lung cancer (NSCLC), pancreatic cancer, renal cell carcinoma (RCC), and melanoma. We conducted our lives-saved and economic valuation analysis for these four cancers.

### Estimation of survival curves

#### Non-small Cell Lung Cancer

First patient recruited in April 2013. ^14^ Overall, there were 13 males and 13 females enrolled in the study where the median age was 63.0 with a range from 40–83 years of age. All patients had stage IV NSCLC. The one-year overall survival rate is 61.5% (95% CI: 42.8%–80.2%) and a two-year overall survival rate of 29.6% (95% CI: 11.6%–47.7%).

We approximated a baseline overall survival rate in the absence of mRNA vaccination by first weighting the age and gender specific overall survival rates across the three types of non-small cell lung cancer reported for 2019. We then weighted these age and gender specific survival rates based on the demographics reported in the study. To obtain the two year survival rate, we first interpolated the age and gender two year survival rate based on the one and three year survival rates from the weighted non-small cell lung cancer. We then computed the weighted average to obtain our baseline estimate for the two year overall survival rate.

We digitized the survival curve to obtain the survival times in order to compute the one and two year survival times with the 95% confidence intervals using the Kaplan Meyer method for the patients treated with the mRNA vaccine.

**Table 1.** One and three year overall survival rates for metastatic non-small cell lung cancer in 2013 ^15,16^.

| Duration | Sex | Age | Type of non-small cell lung cancer |  |  |
| --- | --- | --- | --- | --- | --- |
|  |  |  | Adenocarcinoma | Large cell | Squamous cell carcinoma |
| 1 yr | Female | Under 50 | 56.1% (95CI: 51.3%–60.6%) | 17.9% (95CI: 6.5%–33.8%) | 30.5% (95CI:18%–44%) |
|  |  | 50-64 | 45.4% (95CI:43.3%–47.4%) | 38.6% (95CI: 26.9%–50.1%) | 34.5% (95CI: 30.1%–39%) |
|  |  | 65 and older | 36.6% (95CI: 35.1%–38.1%) | 20.7% (95CI: 12.9%–29.8%) | 29.2% (95CI: 26.7%–31.7%) |
|  | Male | Under 50 | 54.5% (95CI: 49.5%–59.3%) | 17.9% (95CI: 6.5%–33.8%) | 28.3% (95CI:19.9 37.2%) |
|  |  | 50-64 | 35.8% (95CI: 33.9%–37.7%) | 23.1% (95CI: 15.8%–31.4%) | 31.7% (95CI:28.9%–34.7%) |
|  |  | 65 and older | 27.6% (95CI: 26.2%–29.1%) | 18.4% (95CI: 12.1%–25.7%) | 29.2% (95CI:27.1%–31.2%) |
| 3yr | Female | Under 50 | 25.8% (95CI: 21.8%–30%) | 16.6% (95CI: 6.1%–31.7%) | 11% (95CI:4%–21.9%) |
|  |  | 50-64 | 18.4% (95CI: 16.8%–20%) | 14.1% (95CI: 6.9%–23.7%) | 11% (95CI:8.2%–14.2%) |
|  |  | 65 and older | 14.5% (95CI: 13.3%–15.7%) | 7.5% (95CI: 3%–14.6%) | 10.8% (95CI:9.1%–12.7%) |
|  | Male | Under 50 | 23% (95CI: 19%–27.4%) | 16.6% (95CI: 6.1%–31.7%) | 10.2% (95CI:5.2%–17%) |
|  |  | 50-64 | 11.5% (95CI: 10.2%–12.8%) | 5.7% (95CI: 2.3%–11.3%) | 10% (95CI:8.2%–12%) |
|  |  | 65 and older | 8.8% (95CI: 7.9%–9.8%) | 4.4% (95CI: 1.7%–9.4%) | 7.2% (95CI:6%–8.5%) |

**Table 2.**
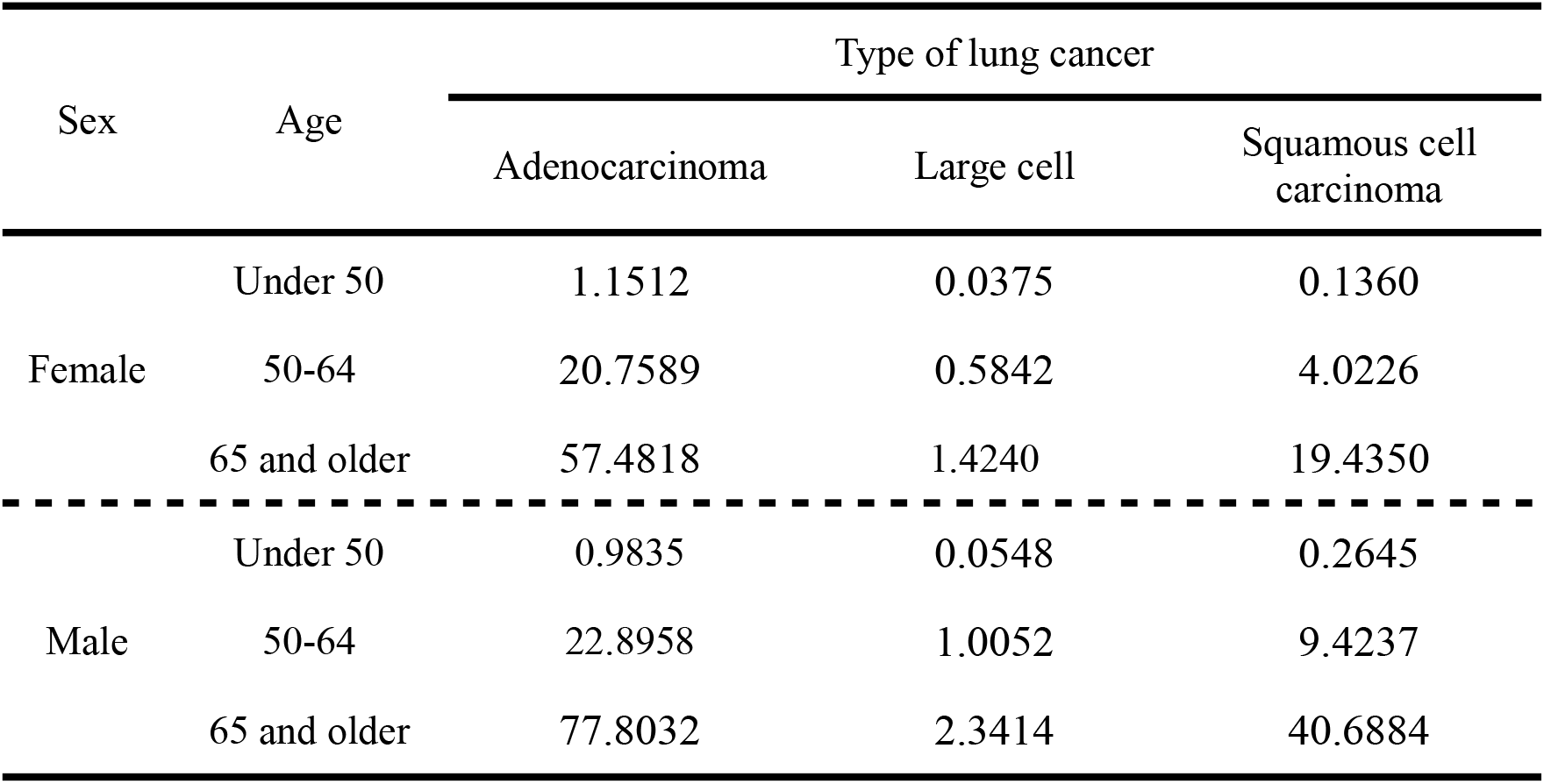
SEER Incidence rates per 100,000 for distant non-scall cell lung cancer in 2013 ^15,16^.

#### Pancreatic Cancer

A total of 16 patients received the mRNA vaccine in combination with other treatment from December 2019 to August 2021.^17^ There were 8 responders and 8 non-responders. The median age among responders was 70.5 years with a range of 59 to 80 years of age, with the median age among non-responders being 71.5 years with a range of 55 to 76. To approximate the proportion of the population under 65, we estimated the parameters of the binomial distribution (chosen since the variance is less than the mean) by fitting 0.005 and 0.995 percentile of the distribution to the reported range and the median to the reported median. From this approximation, we estimate that 10.6% of the population was under the age of 65 years for responders and 17.0% for non-responders.

In the context of the sex of the patients, 6 were female and 2 were male among responders and among non-responders 2 were female and 6 were males. In the context of the stage of cancer for responders, there were 4 patients in stage I (localized); 3 in stage II (regional); and 1 in stage III (regional). For non-responders, there was 1 patient in stage I (localized); 4 in stage II (regional); and 3 in stage III (regional). The treated population was 100% white.

We digitized the survival curve to obtain the survival times in order to compute the one and three year survival times with the 95% confidence intervals for the patients treated with the mRNA vaccine. From the curves provided, we were able to attain the survival times of the unvaccinated patients in the safety-evaluable cohort. We combined responders and non-responders to compute the one and three years overall survival rates among those in the treatment group using the Kaplan-Meier method. The one-year overall survival rate was 100% and the three-year overall survival rate was 75% (95% CI: 53.8%–96.2%). To estimate the lower-bound of the one-year survival rate, we computed the value of *p* that satisfies

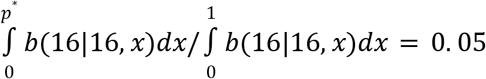

We estimated the lower bound of the confidence interval for the the one-year overall survival rate is *p* = 0. 838. Thus, the one-year overall survival rate was 100% (95% CI: 83.8% –100%).

We approximated a baseline overall survival rate in the absence of mRNA vaccination by weighting the overall survival rates among a white population (includes hispanics) across different ages and stages of cancer reported for 2019 based on the demographics reported in the study.

**Table 3:**
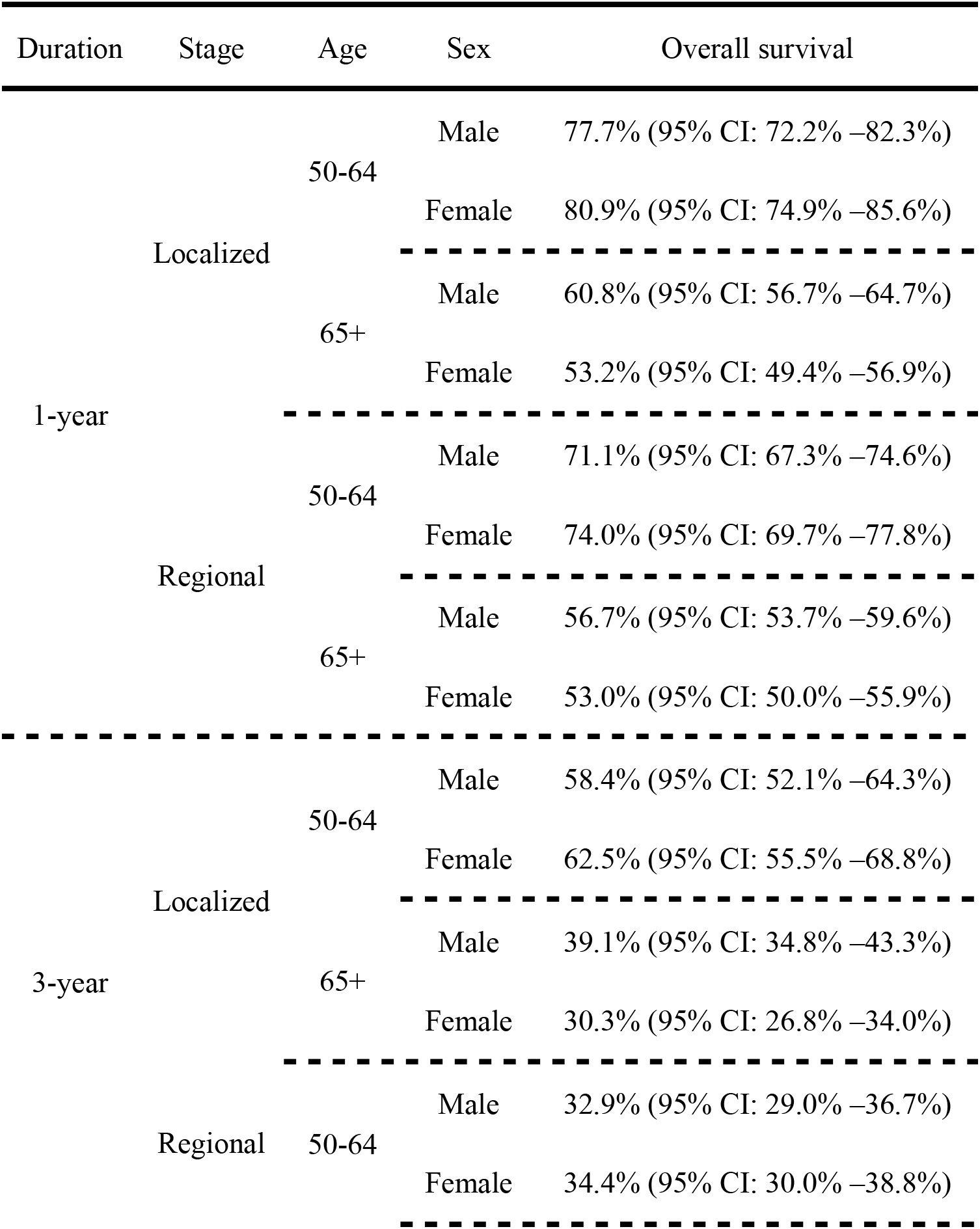
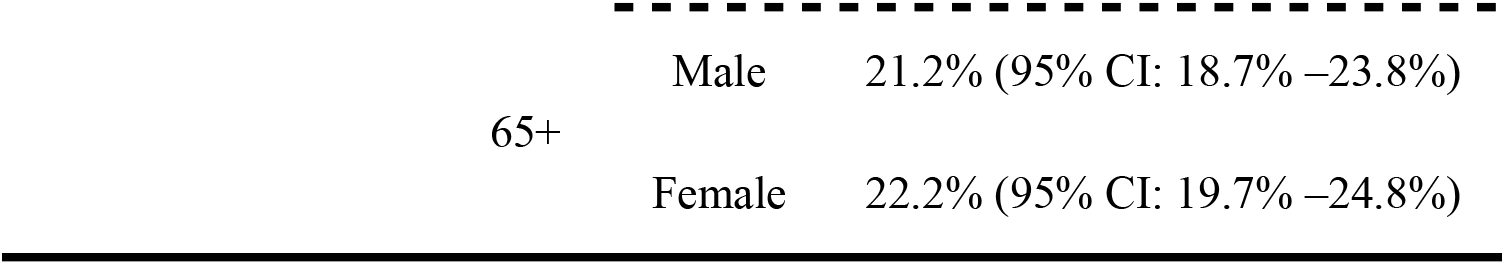
The one and three year overall survival rates among a white population (includes hispanics) for pancreatic cancer for (2019) ^18^.

| Duration | Stage | Age | Sex | Overall survival |
| --- | --- | --- | --- | --- |
| 1-year | Localized | 50-64 | Male | 77.7% (95% CI: 72.2% –82.3%) |
|  |  |  | Female | 80.9% (95% CI: 74.9% –85.6%) |
|  |  | 65+ | Male | 60.8% (95% CI: 56.7% –64.7%) |
|  |  |  | Female | 53.2% (95% CI: 49.4% –56.9%) |
|  | Regional | 50-64 | Male | 71.1% (95% CI: 67.3% –74.6%) |
|  |  |  | Female | 74.0% (95% CI: 69.7% –77.8%) |
|  |  | 65+ | Male | 56.7% (95% CI: 53.7% –59.6%) |
|  |  |  | Female | 53.0% (95% CI: 50.0% –55.9%) |
| 3-year | Localized | 50-64 | Male | 58.4% (95% CI: 52.1% –64.3%) |
|  |  |  | Female | 62.5% (95% CI: 55.5% –68.8%) |
|  |  | 65+ | Male | 39.1% (95% CI: 34.8% –43.3%) |
|  |  |  | Female | 30.3% (95% CI: 26.8% –34.0%) |
|  | Regional | 50-64 | Male | 32.9% (95% CI: 29.0% –36.7%) |
|  |  |  | Female | 34.4% (95% CI: 30.0% –38.8%) |

|  |  |  |
| --- | --- | --- |
| 65+ | Male | 21.2% (95% CI: 18.7% –23.8%) |
|  | Female | 22.2% (95% CI: 19.7% –24.8%) |

#### Renal Cell Carcinoma

The study was conducted among two cohorts with cohort A between August 2003 to June 2004 and cohort B from August 2004 to November 2005. Among cohort A, the mean age of patients was 64.4 years and a range of 36 to 79 years of age and cohort B had an average age of 62.6 years with a range of 44 to 73 years. ^19^ In cohort A 79% (11/14) were male and in cohort B 69% (11/16) were male. Thus, 73.3% (22/30) of the patients treated with an mRNA vaccine were male. All patients were in Stage IV RCC.

To estimate the proportion of the population under the age of 50, between 50 to 64 years of age, and 65 and older, we parameterized a negative binomial fixing its average to the average age reports for the cohort.

We approximated a baseline overall survival rate in the absence of mRNA vaccination across different ages and gender reported for 2004 based on the demographics reported in the study.

We digitized the survival curve to obtain the survival times in order to compute the one, three, five, and ten year survival times with the 95% confidence intervals using the Kaplan Meyer method for the patients treated with the mRNA vaccine.

**Table 4.**
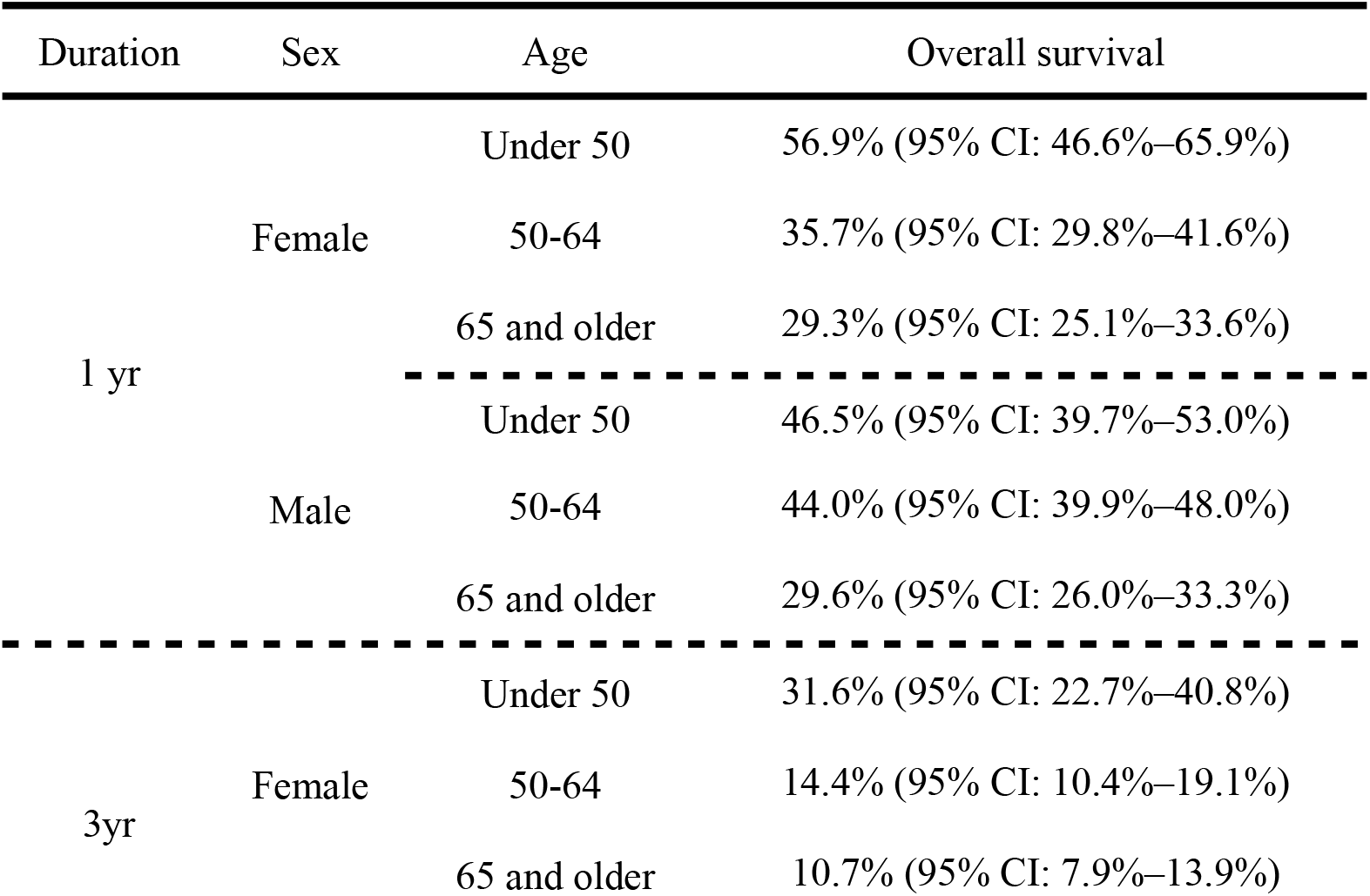
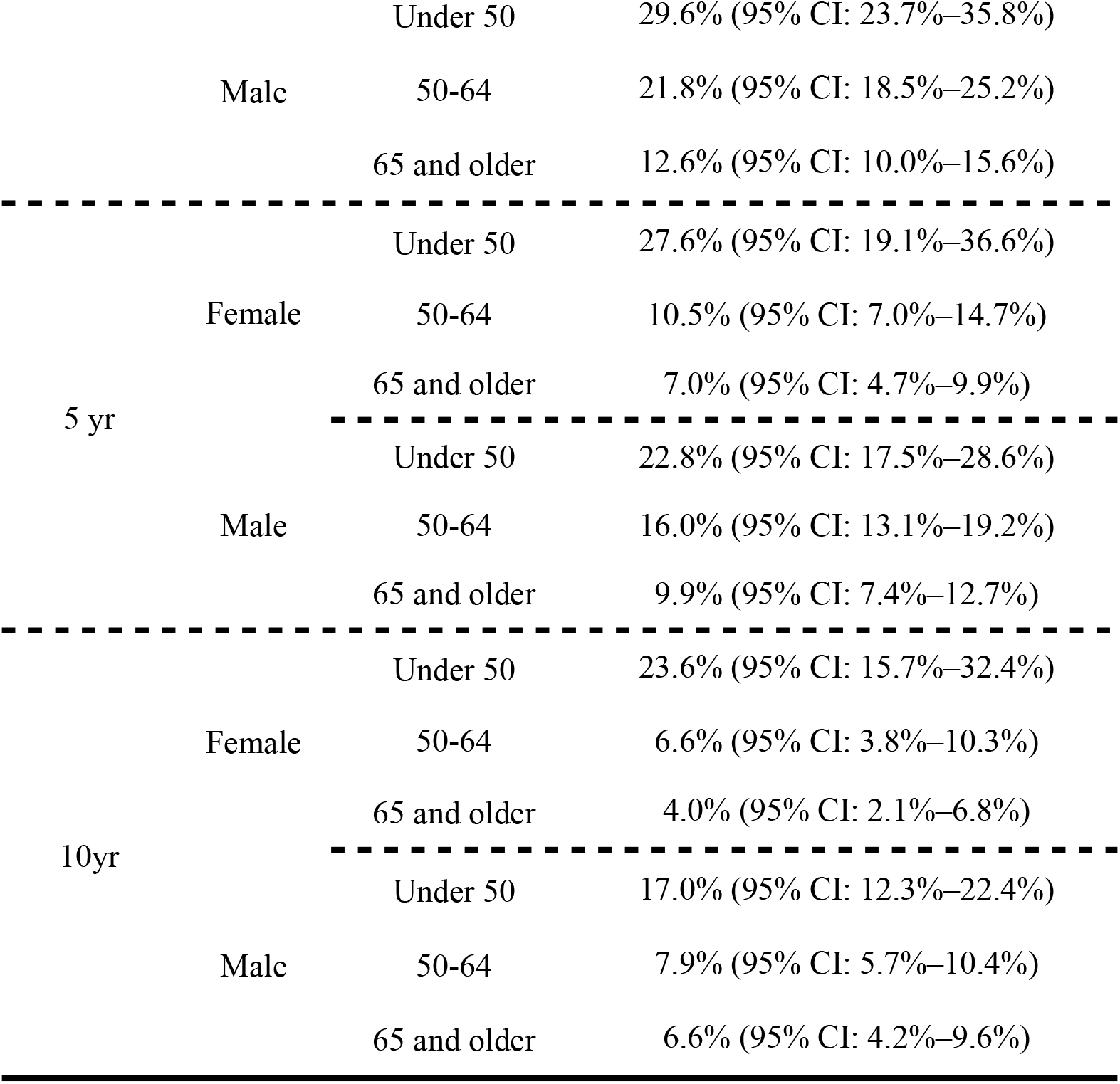
Overall survival rates for distant kidney and renal pelvis cancer for 2004.

#### Melanoma

The use of mRNA-4157 (V940) in combination with KEYTRUDA had a 2.5-year overall survival rate of 96.0% compared to the 90.2% 2.5-year overall survival rate using KEYTRUDA alone. ^21^

The 95% CI for the survival rates were not provided for either group. There were 107 patients who had received the combination treatment of mRNA-4157 and KEYTRUDA and 50 who received the monotherapy of KEYTRUDA ^22^. Using the beta distributions

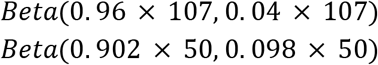

we estimate the 95% confidence interval for mRNA-4157 (V940) in combination with KEYTRUDA to be (0.9157–0.9882) and (0.8067–0.9672) for the patients under the KEYTRUDA monotherapy treatment.

Within the vaccine treated group 65% (70/107) were male, 55.1% (59/107) were under the age of 65 with a median age of 63 and IQR of 53–72, and 85% (91/107) were in stage III and 15% (16/107) in stage IV. We estimated the parameters of a negative binomial distribution to the percentiles reported Using this parameterized distribution, we estimated that 31.1% of the population under 65 consisted of individuals under the age of 50. Thus, 17.2% of the population is estimated to be under the age of 50 and 37.9% between the ages of 50 and 64 years of age

Using the 1-year and 3 year survival rates from 2019 for population group *j*, we interpolated the 2.5 year survival rate. Then we computed the weighted average of the 2.5 year survival rate based on the population demographics described in the study.

**Table 5:**
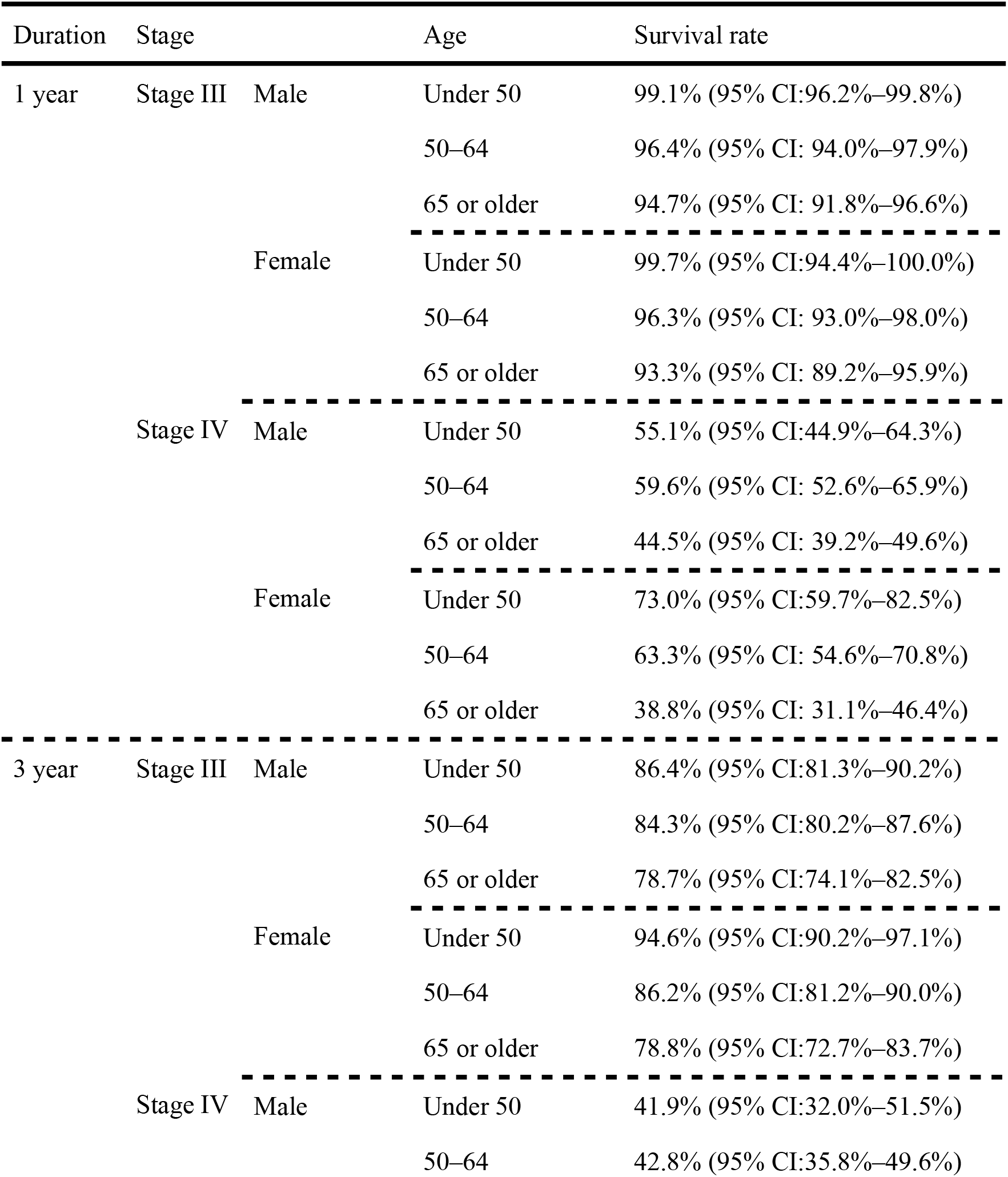
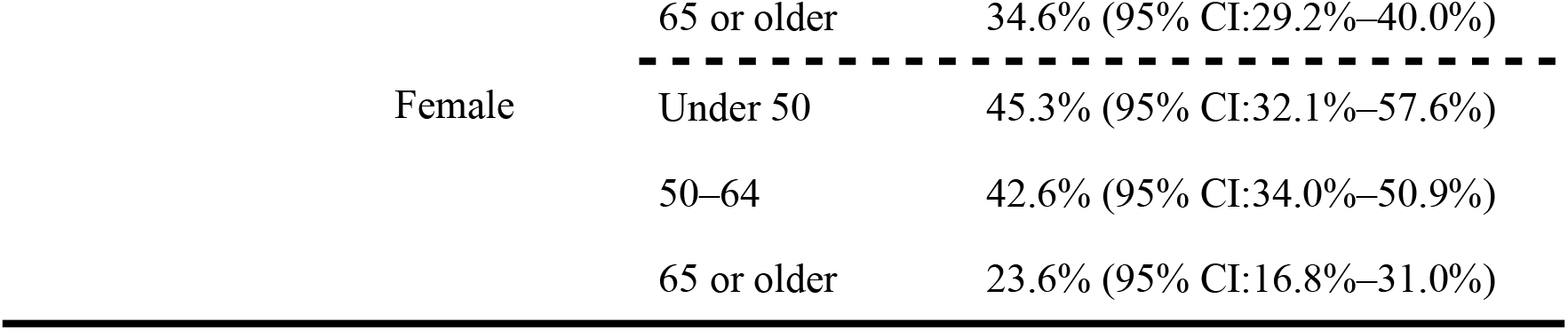
One and three year overall survival rates for melanoma reported for 2019.^23^.

| Duration | Stage |  | Age | Survival rate |
| --- | --- | --- | --- | --- |
| 1 year | Stage III | Male | Under 50 | 99.1% (95% CI:96.2%–99.8%) |
|  |  |  | 50–64 | 96.4% (95% CI: 94.0%–97.9%) |
|  |  |  | 65 or older | 94.7% (95% CI: 91.8%–96.6%) |
|  |  | Female | Under 50 | 99.7% (95% CI:94.4%–100.0%) |
|  |  |  | 50–64 | 96.3% (95% CI: 93.0%–98.0%) |
|  |  |  | 65 or older | 93.3% (95% CI: 89.2%–95.9%) |
|  | Stage IV | Male | Under 50 | 55.1% (95% CI:44.9%–64.3%) |
|  |  |  | 50–64 | 59.6% (95% CI: 52.6%–65.9%) |
|  |  |  | 65 or older | 44.5% (95% CI: 39.2%–49.6%) |
|  |  | Female | Under 50 | 73.0% (95% CI:59.7%–82.5%) |
|  |  |  | 50–64 | 63.3% (95% CI: 54.6%–70.8%) |
|  |  |  | 65 or older | 38.8% (95% CI: 31.1%–46.4%) |
| 3 year | Stage III | Male | Under 50 | 86.4% (95% CI:81.3%–90.2%) |
|  |  |  | 50–64 | 84.3% (95% CI:80.2%–87.6%) |
|  |  |  | 65 or older | 78.7% (95% CI:74.1%–82.5%) |
|  |  | Female | Under 50 | 94.6% (95% CI:90.2%–97.1%) |
|  |  |  | 50–64 | 86.2% (95% CI:81.2%–90.0%) |
|  |  |  | 65 or older | 78.8% (95% CI:72.7%–83.7%) |
|  | Stage IV | Male | Under 50 | 41.9% (95% CI:32.0%–51.5%) |
|  |  |  | 50–64 | 42.8% (95% CI:35.8%–49.6%) |

|  |  |  |
| --- | --- | --- |
|  | 65 or older | 34.6% (95% CI:29.2%–40.0%) |
| Female | Under 50 | 45.3% (95% CI:32.1%–57.6%) |
|  | 50–64 | 42.6% (95% CI:34.0%–50.9%) |
|  | 65 or older | 23.6% (95% CI:16.8%–31.0%) |

### Effects of mRNA vaccination on the overall survival rates

To estimate the baseline overall survival rates reflective of the demographics of the population in the published study, we weighted SEER one and three year overall survival rates for a year similar to the study period. For studies which had a control group, we also utilized the survival rates from this group as a representative of baseline overall survival rates in the absence of mRNA vaccination.

For each of the survival rates with and without mRNA vaccination, we parameterized a beta distribution such that the mode was fixed to the reported point estimate and fitting the 2.5^th^ and 97.5^th^ percentiles to the reported 95% confidence intervals. For studies not having the one or three year overall survival rates for the mRNA vaccine, we extrapolated the values to approximate the survival rate at the missing year.

Using the estimates of the one year overall survival rates, we evaluated the benefit of mRNA vaccination on increasing survival rates through a logistical regression model

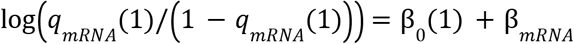

where *q*_0_ (1) = 1/(1 + exp_{_− β_0_(1)}) is the one year survival rate without mRNA vaccination.

Similarly, using the estimates of the three year overall survival rates, we evaluated the benefit of mRNA vaccination on increasing survival rates through a logistical regression model

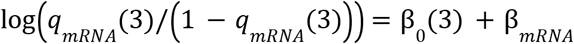

where *q*_0_ (3) = 1/(1 + exp_{_− β_0_ (3)}) is the three year survival rate without mRNA vaccination.

We estimated the parameters for the logistic regression through maximizing the log-likelihood

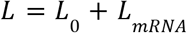

where the log-likelihood for the one and three year survival rate without mRNA is

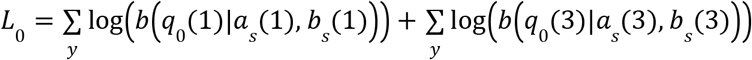

and the log-likelihood for the one year survival rate with mRNA is

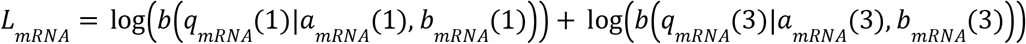

where *b*(*x*|*a, b*) is the Beta probability density function and *y* is the number of estimates for the one year overall survival rate without mRNA vaccination. We used Bayesian melding to construct uncertainty for estimate of β_*mRNA*_.

Using recent (2021) age and stage specific one year survival rates for the specified cancer, we estimated the one year survival rate for those receiving mRNA vaccination treatment by

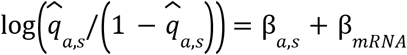

where 1/( 1 + exp{ − β_*a,s*_}) is the age and stage specific one year survival rate reported for 2021 in the SEER dataset. We used a similar approach for the three year survival rates (reported or 2019), assuming the effect of mRNA vaccination was the same as the first year.

#### Cost

Given death occurs within three years since diagnosis,if *t* ∈ [0, 1] then the cumulative probability function for death is expressed as

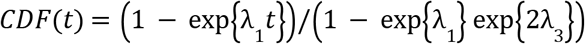

and if *t* ∈ [1, 3] then the cumulative probability function for death is expressed as

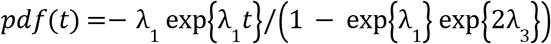

where λ_1_ = log (σ _*c,a,s*_ (1)) and λ_3_ = log(σ _*c,a,s*_ (3)/σ_*c,a,s*_ (1)) /2 with σ_*c,a,s*_ (1) being the on year survival rate and σ_*c,a,s*_ (3) the three-year survival rate.

If *t* ∈ [0, 1] then the probability density function is

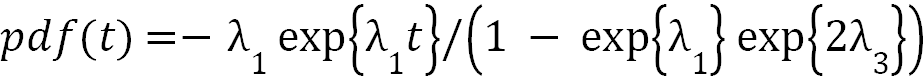

and *t* ∈ [1, 3] then the probability density function is

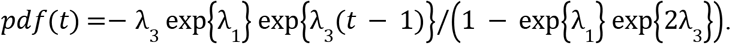

Given an individual dies from cancer within three years since diagnosis, we computed the average time from diagnosis to death

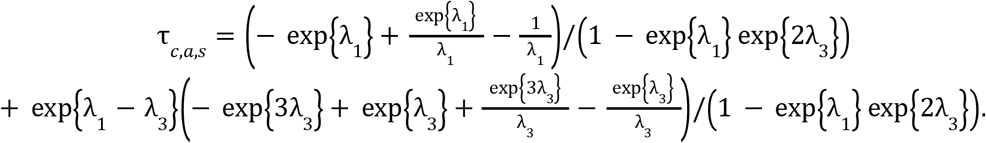

We computed this value for those treated with mRNA vaccination, denoted 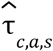, and those who were not τ_*c,a,s*_.

For those receiving treatment and not surviving in the average time of extending life is

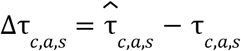

and among the averted deaths over the three year period the average time of extending life is

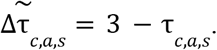

Using a 3% discounting rate, *r* = 0. 03, we computed the life years gained

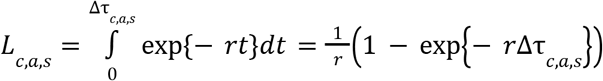

and

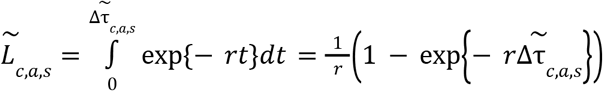

Using $604,000for the central value of a statistical life year, the cost associated with the life years lost is expressed as

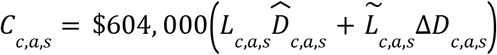

where Δ*D*_*c,a,s*_ is the number of deaths averted over the three year period due to mRNA vaccination and 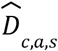 is the number of cancer related deaths over the three year period among those treated with an mRNA vaccine.

These costs among each age, sex, and stage of cancer were then aggregated to obtain the overall costs associated with the life years lost in the absence of mRNA vaccination.

## Results

We identified 32 active mRNA cancer vaccine clinical trials currently underway in the United States, including 22 classic mRNA vaccines and 10 dendritic cell (DC) mRNA vaccines. Among classic mRNA vaccines, three trials are in Phase III or Phase II/III, fourteen are in Phase II or Phase I/II, and five are in Phase I. Among DC-mRNA vaccines, four are in Phase II or Phase I/II, and six are in Phase I. The three most advanced trials (Phase III or Phase II/III) are projected to complete in 2027, 2029, and 2035, respectively. Because most DC-mRNA vaccines remain early-phase with limited published results, they were not included in our primary analysis. Using improvements in overall survival, recurrence-free survival, and disease progression as indicators of clinical promise, we designated 11 trials as particularly promising. These studies focus on malignant melanoma, non-small cell lung cancer (NSCLC), pancreatic cancer, bladder cancer, colorectal cancer, and cutaneous squamous cell carcinoma (**Table 6**). Trial characteristics, including phase and estimated completion, were obtained from ClinicalTrials.gov.

**Table 6.** mRNA-Vaccine Cancer Treatments in Clinical Trials in the United States demonstrating promising results.

| **Cancer Type** | **Vaccines** | **Current Trial Phase** | **Trial ID** | **Estimated Study Completion** |
| --- | --- | --- | --- | --- |
| Malignant Melanoma | mRNA-4157 vaccine/neoantigen + Keytruda | Phase III | NCT05933577 | 2029-09-09 |
|  | BNT111 TYR vaccine | Phase II | NCT04526899 | 2025-12 |
|  | BNT122 (autogene cevumeran) vaccine | Phase II | NCT03815058 | COMPLETED 2025-01-21 |
|  | mRNA-4157 vaccine/neoantigen + Keytruda | Phase II | NCT03897881 | 2029-09-09 |
| Non-Small Cell Lung Cancer | mRNA-4157 vaccine/neoantigen + Keytruda | Phase III | NCT06077760 | 2035-12-21 |
|  | mRNA Vaccine [BI 1361849] | Phase I/II | NCT03164772 | COMPLETED 2021-10-29 |
| Pancreatic Cancer | BNT122 (autogene cevumeran) vaccine | Phase II | NCT05968326 | 2029-12-27 |
|  | mRNA lipoplex vaccine with FOLFIRINOX (chemotherapy) | Phase I | NCT04161755 | 2025-11-11 |
| Bladder Cancer | mRNA-4157 vaccine/neoantigen + Keytruda | Phase I/II | NCT06305767 | 2031-10-20 |
| Colorectal Cancer | GRANITE (GRT-C901/ GRT-R902) vaccine | Phase II/III | NCT05141721 | 2027-03 |
| Cutaneous Squamous Cell Carcinoma | mRNA-4157 vaccine/neoantigen + Keytruda | Phase II | NCT06295809 | 2026-03-05 |

Two major development programs dominate this landscape. Moderna and Merck are jointly evaluating the mRNA-4157 neoantigen vaccine in combination with pembrolizumab (Keytruda), with five ongoing trials included among our promising set. Results from trial NCT03897881 showed that mRNA-4157 (V940) plus Keytruda reduced the risk of recurrence or death by 49% and the risk of distant metastasis or death by 62% compared with Keytruda alone in malignant melanoma, providing strong justification for expansion into additional cancer types. In parallel, BioNTech and Genentech are advancing BNT122 (autogene cevumeran), which is being tested alongside chemotherapy across multiple malignancies. Two of these trials are included as promising based on encouraging immunogenicity and delayed recurrence signals.

Within malignant melanoma, the combination of mRNA-4157 and Keytruda has demonstrated clinically significant improvements in both recurrence-free survival and distant metastasis-free survival, supporting five active trials of this regimen. In NSCLC, one Phase III and one Phase I/II trial are ongoing, both reporting improved one- and three-year survival compared with standard therapies. For pancreatic cancer, a Phase I trial completing in 2025 has shown encouraging preliminary results, while a Phase II study has suggested delayed recurrence and durable immune responses. In bladder cancer, a Phase II mRNA-4157 trial is advancing, supported by efficacy signals from parallel melanoma studies. In colorectal cancer, a Phase II/III study expected to complete in 2027 has reported reduced disease progression in low-burden patient groups. Finally, in cutaneous squamous cell carcinoma, a Phase II mRNA-4157 trial, projected to complete in 2026, has already demonstrated improvements in one- and three-year survival rates.

### Cancer deaths averted through mRNA vaccination treatment

Based on available studies that have reported overall survival rates with mRNA vaccines, which exhibit a promising increase in overall survival for cancer patients (Table 7), we focused our analysis on non-small cell lung cancer, pancreatic cancer, renal cell cancer, and melanoma. Assuming all diagnosed patients received mRNA vaccination, we estimated the number of deaths averted among these four cancers compared to no mRNA vaccination within a single annual cohort of patients (**Figure 1**).

**Table 7.**
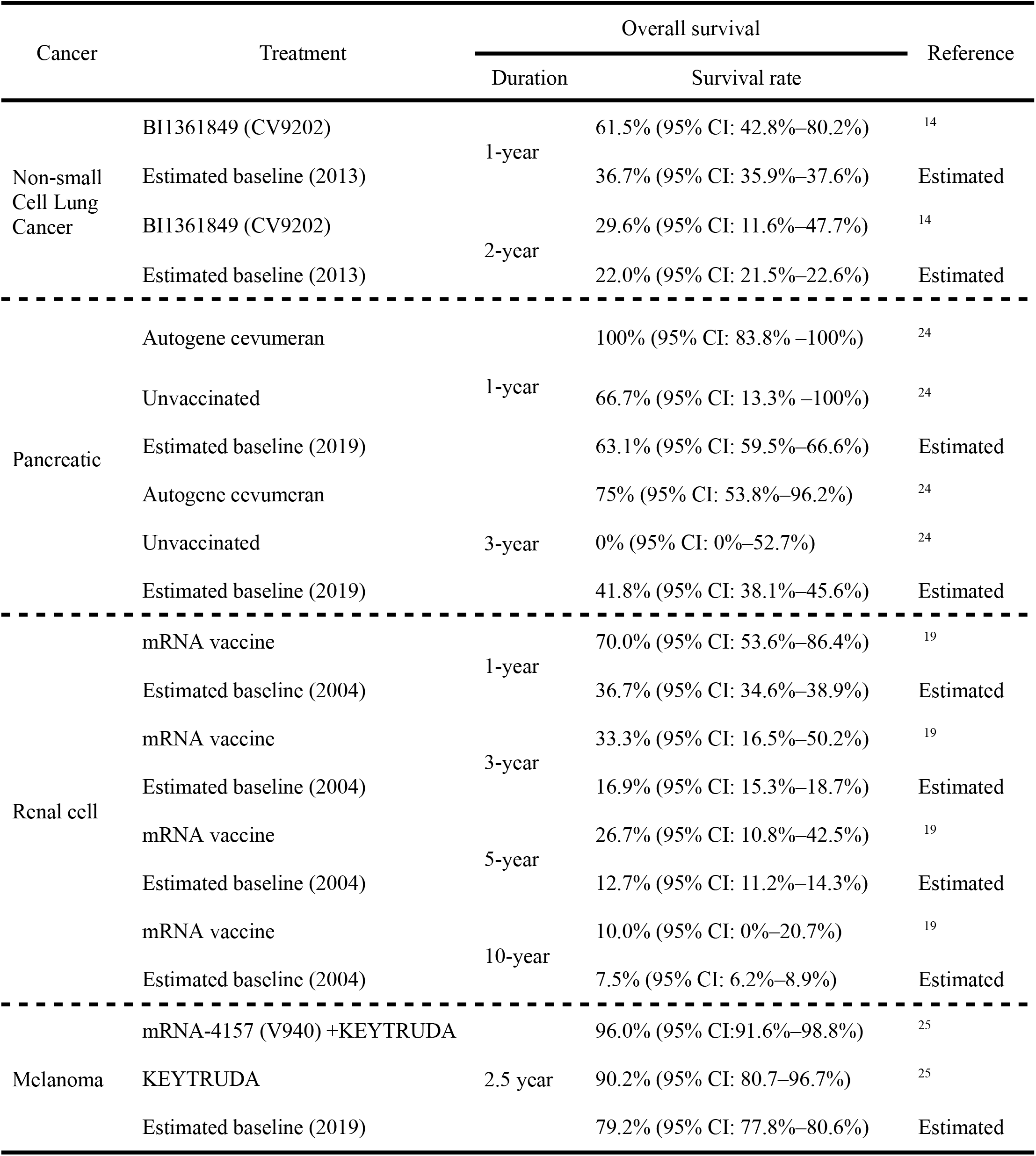
The overall survival rate for non-small cell lung cancer, pancreatic cancer, renal cell cancer, and melanoma under treatment with and without mRNA vaccination.

**Figure 1:**
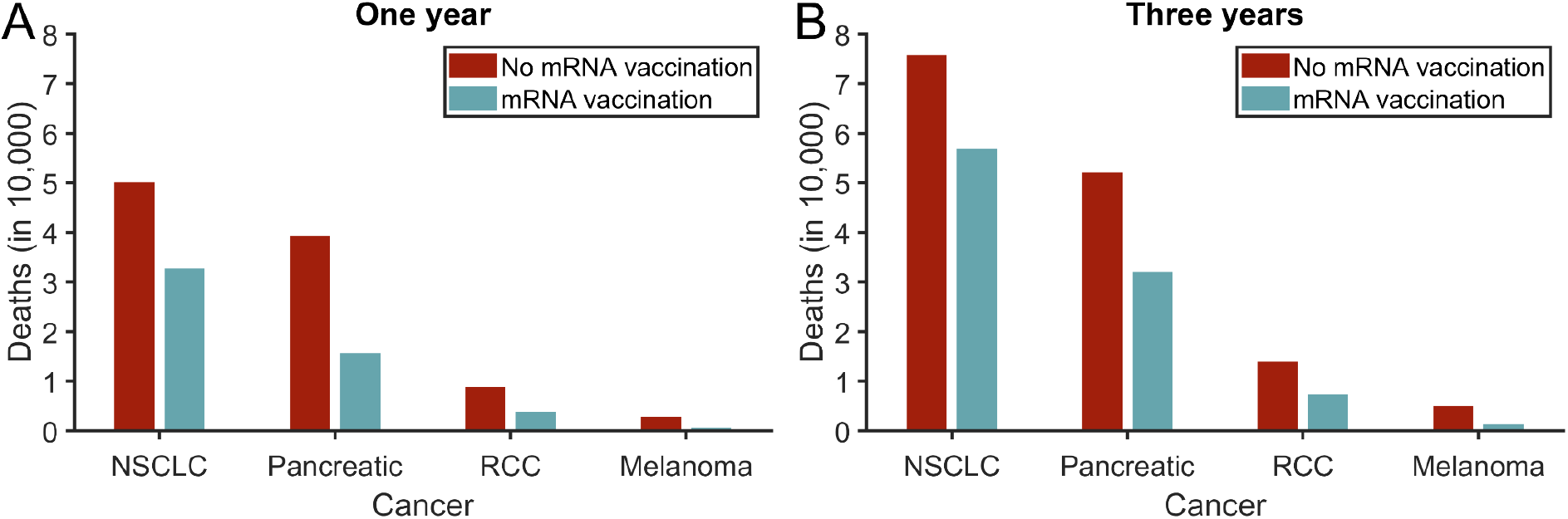
The estimated number of deaths for non-small cell lung cancer (NSCLC), pancreatic cancer, renal cell cancer (RCC), and melanoma with no mRNA vaccination treatment (red) and with mRNA vaccination treatment (green) over a span of (A) one year since diagnosis and (B) three years since diagnosis.

#### Non-small cell lung cancer

Using age and gender specific rates for non-small cell lung cancer (i.e., adenocarcinoma of the lung and bronchus, large cell carcinoma of the lung and bronchus, as well as squamous cell carcinoma of the lung and bronchus), we estimated an annual incidence of 136792 (95% CrI: 136041–137553). Among these cases, we estimated there to be 50077 (95% CrI: 49762–50389) deaths within one year and 75780 (95% CrI: 75348–76222) deaths within three years since diagnosis in the absence of mRNA vaccination. We estimate that the use of mRNA vaccination to treat non-small cell lung cancer could avert upwards to 17274 (95% CrI: 1261–29635) over one year and 18947 (95% CrI: 1280–35402) over the span of three years.

#### Pancreatic

Using age and gender specific rates for pancreatic cancer, we estimated an annual incidence of 62999 (95% CrI: 62570–63443). Among these cases, we estimated there to be 39196 (95% CrI: 38921–39528) deaths within one year and 52072 (95% CrI: 51700–52458) deaths within three years since diagnosis in the absence of mRNA vaccination. We estimate that the use of mRNA vaccination to treat pancreatic cancer could avert upwards to 23563 (95% CrI: 12527–32925) over one year and 20075 (95% CrI: 9209–34033) over the span of three years.

#### Renal cell cancer

Using age and gender specific rates for renal cell cancer (i.e., renal pelvis and kidney), we estimated an annual incidence of 77724 (95% CrI: 77216–78227). Among these cases, we estimated there to be 8839 (95% CrI: 8735–8947) deaths within one year and 13949 (95% CrI: 13805–14088) deaths within three years since diagnosis in the absence of mRNA vaccination. We estimate that the use of mRNA vaccination to treat renal cell cancer could avert upwards to 4921 (95% CrI: 3054–6358) over one year and 6613 (95% CrI: 3962–8896) over the span of three years.

#### Melanoma

Using age and gender specific rates for melanoma, we estimated an annual incidence of 95215 (95% CrI: 94619–95832). Among these cases, we estimated there to be 2840 (95% CrI: 2791–2890) deaths within one year and 5008 (95% CrI: 4923–5116) deaths within three years since diagnosis in the absence of mRNA vaccination. We estimate that the use of mRNA vaccination to treat melanoma could avert upwards to 2235 (95% CrI: 1800–2499) over one year and 3691 (95% CrI: 2962–4266) over the span of three years.

### Economic cost of years of life lost

To quantify the economic cost to the United States attributed to the cessation of the development of mRNA vaccination, we applied the central estimate of $604,000 for the value per statistical life year to the years of life lost in the absence of mRNA vaccination in a single cohort (based on annual incidence) up to three years from the time of diagnosis using a 3% discounting rate ^26^.

We estimate that among a single cohort of non-small cell lung cancer, pancreatic cancer, renal cell cancer, and melanoma patients, ceasing government investment in the mRNA platform could cost the United States an additional $75.44 (95% CrI: $44.18–$103.32) billion over a three year period.

#### Non-small cell lung cancer

The estimated costs associated with the lives lost in a single cohort over a three year period due to the absence of mRNA vaccination treatment is $27.85 (95% CrI: $1.99–$49.20) billion.

#### Pancreatic

The estimated costs associated with the lives lost in a single cohort over a three year period due to the absence of mRNA vaccination treatment is $34.93 (95% CrI: $17.92–$52.06) billion.

#### Renal cell cancer

The estimated costs associated with the lives lost in a single cohort over a three year period due to the absence of mRNA vaccination treatment is $8.50 (95% CrI: $5.20–$11.16) billion.

#### Melanoma

The estimated costs associated with the lives lost in a single cohort over a three year period due to the absence of mRNA vaccination treatment is $4.15 (95% CrI: $3.37–$4.75) billion.

## Discussion

While prior work has highlighted the promise of mRNA technology in oncology, no analysis has yet translated early clinical results into projected lives saved or monetized these gains using nationally representative cancer incidence and survival data. By combining preliminary trial outcomes with U.S. cancer registry statistics and standard federal economic valuation metrics, we provide an evidence-based estimate of what is at stake if federal investment in mRNA vaccine development is curtailed. The therapeutic progress demonstrated by each of the clinical trials in our analysis has the potential to avert nearly 50,000 deaths with an economic value of $75 billion. These estimates represent only a single annual cohort of patients treated for their respective cancer. Disinvestment in mRNA oncology research would cause an accumulation of averted deaths and associated costs.

The repercussions of curtailing funding for mRNA vaccine development will be far-reaching. Beyond oncology, mRNA vaccines hold strong promise for the treatment of infectious, metabolic, autoimmune, genetic, and chronic diseases and disorders ^27,28^. Future development of mRNA may also include pandemic preparedness efforts, including mRNA vaccine libraries for known pathogens ^27,28^. The ability of mRNA vaccines to be redesigned quickly for new variants or novel pathogens, scaled rapidly to millions of doses, and adapted for diverse targets (such as influenza, HIV, or EBV) positions mRNA vaccines as a cornerstone of infectious-disease control. Continued government investment would therefore function not only as healthcare spending but as a high-return insurance policy against future crises.

A loss of funding for mRNA vaccine development will both impact existing clinical trials and hinder future innovation. Moreover, this disinvestment may undermine public trust in vaccines and mRNA technology, which has the potential to continue to be a critical biomedical innovation ^28,29^. Given mRNA vaccines treat a spectrum of diseases and the burden of cancer is pervasive across demographics, investment in mRNA technology is crucial to ensure the health of all.

## Data Availability

All data produced in the present work are contained in the manuscript.

